# Kinetics of viral load, immunological mediators and characterization of a SARS-CoV-2 isolate in mild COVID-19 patients during acute phase of infection

**DOI:** 10.1101/2020.11.05.20226621

**Authors:** Anbalagan Anantharaj, Sunil Gujjar, Saurabh Kumar, Nikhil Verma, Jigme Wangchuk, Naseem Ahmed Khan, Aleksha Panwar, Akshay Kanakan, Vivekanand A, Janani Srinivasa Vasudevan, Asim Das, Anil Kumar Pandey, Rajesh Pandey, Guruprasad R. Medigeshi

**Affiliations:** Clinical and Cellular Virology lab and Bioassay Laboratory, Translational Health Science and Technology Institute, Faridabad, Haryana. INDIA; Employees State Insurance Corporation Medical College and Hospital, Faridabad, Haryana, INDIA; CSIR-Institute of Genomics and Integrative Biology, Delhi, INDIA; Academy of Scientific and Innovative Research (AcSIR), Ghaziabad-201002, INDIA

**Keywords:** SARS-CoV-2, COVID-19, Cytokines, Chemokines, Inflammation, Zinc, epithelial cells

## Abstract

Over 95% of the COVID-19 cases are mild-to-asymptomatic who contribute to disease transmission whereas most of the severe manifestations of the disease are observed in elderly and in patients with comorbidities and dysregulation of immune response has been implicated in severe clinical outcomes. However, it is unclear whether asymptomatic or mild infections are due to low viral load or lack of inflammation. We have measured the kinetics of SARS-CoV-2 viral load in the respiratory samples and serum markers of inflammation in hospitalized COVID-19 patients with mild symptoms. We observed a bi-phasic pattern of virus load which was eventually cleared in most patients at the time of discharge. Viral load in saliva samples from a subset of patients showed good correlation with nasopharyngeal samples. Serum interferon levels were downregulated during early stages of infection but peaked at later stages correlating with elevated levels of T-cell cytokines and other inflammatory mediators such as IL-6 and TNF- α which showed a bi-phasic pattern. The clinical recovery of patients correlated with decrease in viral load and increase in interferons and other cytokines which indicates an effective innate and adaptive immune function in mild infections. We further characterized one of the SARS-CoV-2 isolate by plaque purification and show that infection of lung epithelial cells (Calu-3) with this isolate led to cytopathic effect disrupting epithelial barrier function and tight junctions. Finally we showed that zinc was capable of inhibiting SARS-CoV-2 infection in this model suggesting a beneficial effect of zinc supplementation in COVID-19 infection.

**IMPORTANCE:** A majority of COVID-19 patients are asymptomatic or exhibit mild symptoms despite high viral loads suggesting a key role for the acute phase innate immune response in limiting the damage and clearing the virus. Therefore, it is important to understand the early phase response to SARS-CoV-2 infection in such patients to devise strategies for clinical management of the disease. Our study shows the kinetics of immune mediators in the serum samples collected from hospitalized COVID-19 patients with mild symptoms. We further characterize a virus isolate from one of these patients and demonstrate its effect on epithelial barrier functions and show that zinc was capable of inhibiting SARS-CoV-2 infection under these conditions. Our results suggest a key role for the innate immune responses in the early phase of infection in mitigating clinical symptoms, clearing the virus and recovery from illness and suggest an antiviral role for zinc in COVID-19 infection.

## INTRODUCTION

Coronavirus disease of 2019 (COVID-19) has infected over 35 million people worldwide leading to over a million deaths (as on 10th October 2020). Severe acute respiratory syndrome coronavirus-2 (SARS-CoV-2), a positive-sense RNA virus from the family *Coronaviridae* is the causative agent of COVID-19. The RNA genome of SARS-CoV-2 ranges from 26 to 32 kilobases in length and similar to other coronaviruses that have jumped species to infect humans, SARS-CoV-2 is believed to have been in circulation in animals such as bats for several decades before infecting humans (1). The genome of SARS-CoV-2 shares about 96% identity with that of bat coronavirus and is about 80% identical to the SARS-CoV-1 (2). Angiotensin converting enzyme-2 (ACE2) has been reported as a primary entry receptor for SARS-CoV-2 (3). ACE2, a type I membrane protein, is expressed in lungs, kidneys, intestine and heart. The spike glycoprotein (S-protein) on the SARS-CoV-2 virion surface mediates the receptor recognition and membrane fusion thus facilitating the entry of virus in the cell (4). There are close to 200 vaccines under development for COVID-19 and some of the vaccines have advanced to Phase III trials (5). Efforts to repurpose drugs and to manage the cytokine storm have identified a number of drugs that have been recommended for use in COVID-19 (6) but an effective antiviral specific to SARS-CoV-2 is yet to be licensed for human use.

Most of the patients with COVID-19 exhibit mild or no symptoms and severe clinical symptoms are observed in patients in the older age group (>65 years) or patients with underlying comorbidities such as hypertension and diabetes. Impaired immune response and cytokine storm has been implicated in clinical manifestations observed in severe disease (7-9). Increased viral load has been shown to correlate with disease severity (7, 10) and some of the antivirals such as Remdesivir which inhibit the viral RNA-dependent-RNA-polymerase of SARS-CoV-1, Middle East Respiratory Syndrome Coronavirus (MERS-CoV) and SARS-CoV-2 has been shown to reduce time to recovery and mortality in COVID-19 patients (11). As is the case with RNA viruses, coronaviruses accumulate mutations and evolve at a rate similar to other RNA viruses (12, 13). Variations in the viral genome and emergence of novel clades has been implicated in increased fitness and transmission (14) however there is no evidence to suggest increased virulence of infecting virus as a cause of severe disease in COVID-19 patients suggesting that the host response and underlying comorbidities plays a key role in clinical outcomes (15). In this study, we monitored the kinetics of viral load in respiratory samples and saliva and cytokines and chemokines in the serum of hospitalized COVID-19 patients who displayed mild/moderate symptoms and recovered from illness. We isolated SARS-CoV-2 from one of the patients and further characterized the isolate by plaque-purification and sequencing and assessed the effect of virus infection on epithelial barrier functions. Our results suggest a clear inverse correlation between interferon response and viral load and identifies signatures in the inflammatory mediators in the serum of young patients who recover from COVID-19. We also show that the virus isolate from a patient with mild infection is capable of infecting lung epithelial cells and disrupting epithelial barrier functions leading to cell death further underscoring the importance of a robust host response in mitigating the clinical outcomes in COVID-19 infection. Finally, addition of zinc post-infection inhibited SARS-CoV-2 infection indicating the antiviral function of zinc in SARS-CoV-2 life-cycle.

## MATERIALS AND METHODS

### Human Ethics

The study was approved by the Institutional ethics committees for human research at both the institutions (No. 134/A/11/16/Academics/MC/2016/134 and THS 1.8.1/ (93)). Informed consent was obtained from all the participants.

### Clinical samples

Patients with symptoms of COVID-19 infection were diagnosed using RealStar® SARS-CoV-2 RT-PCR kit (altona Diagnostics GmbH) at ESIC Medical College & Hospital. Clinical presentations were mild or asymptomatic (identified by contact tracing) and included mild to moderate fever, dry cough, and loss of sense of smell and taste. As per the regulations at the time of the study, all patients with a positive RT-PCR test were admitted to the hospital within 24-48 hours of testing positive and treated with azithromycin with hydroxychloroqine, Vitamin C and D. Samples were collected from patients who consented to participate in the study by signing the informed consent form. Nasopharyngeal/oropharyngeal (NP/OP) swab in virus transport medium (VTM), saliva and serum was collected from Day 1 of enrolment till discharge. Saliva was collected in specimen collection cups early in the morning immediately after the patient woke up from sleep. All the samples collected daily at same interval. All patients were isolated in the hospital for at least 14 days.

### Virus culture and isolation

SARS-CoV-2 Isolate USA-WA1/2020 was obtained from BEI resources (deposited by the Centers for Disease Control and Prevention and obtained through BEI Resources, NIAID, NIH: SARS-Related Coronavirus 2, Isolate USA-WA1/2020, NR-52281). Indian isolates of SARS-CoV-2 was isolated from COVID-19-positive VTM samples (VTM) using Vero E6 cells (Obtained from National Centre for Cell Science, Pune, INDIA). 2×10^6^ cells were grown in a T25 flask to about 80% confluency. Culture medium was removed and washed once with 5 ml of phosphate-buffered saline (PBS). 500 μl of VTM was added onto the cells and the flask was incubated for 1 h at 37°C in a CO_2_ incubator with intermittent mixing. After 1h of adsorption, monolayer was washed twice with 1X PBS and DMEM supplemented with 2% fetal bovine serum (FBS) and 2X antibiotics (200 units of penicillin, 200 μg of streptomycin) and 250 ng/ml amphotericin B and non-essential amino acids. Cell monolayer was observed under the microscope every day. Cytopathic effect was observed usually by day two of infection when the Ct value of the sample was less than 20 in RT-PCR. Culture supernatant was harvested and clarified at 1000 x *g* and multiple small aliquots were prepared and stored at −80°C.Virus titer in the supernatants were estimated by plaque assays on Vero E6 cells as per the previous protocols (16) except that the cells were fixed at 60 h pi. Clinical specimens and virus cultures were handled in biosafety level 3 (BSL3) facility as per the standard operating procedure and guidelines for BSL3 level pathogen.

### Quantitative RT PCR

RNA was extracted from 150 μl of VTM using Viral RNA Isolation Kit (MACHEREY-NAGEL), and eluted in 50 μl of nuclease-free water and used as a template for quantitation of SARS-CoV-2 viral RNA levels by reverse transcription polymerase chain reaction (RT-PCR). 2019-nCoV CDC Probe and Primer kit for SARS-CoV-2 manufactured (Biosearch Technologies) was used to detect N gene by real time RT PCR. RNAse P was detected as endogenous control for normalization. The region of N gene starting from 28287 – 29230 was amplified using the forward primer 5’-GACCCCAAAATCAGCGAAAT-3’ and the reverse primer 5’-GCGCGACATTCCGAAGAA-3’ and cloned into pGEM®-T-Easy vector (Promega). This clone was linearized using Sac II enzyme and *in vitro* transcribed using the SP6 RNA polymerase (Promega). The transcript was purified and used as a template for generating standard curve to estimate the copy number.

For virus isolates, RNA was isolated from 4 different plaque-purified isolates using QIAamp Viral RNA mini kit (Qiagen) according to manufactures instruction. SARS-CoV-2 viral RNA was detected using RealStar® SARS-CoV-2 RT-PCR kit (altona Diagnostics GmbH). To check the presence of other respiratory virus in samples, FTlyo respiratory pathogen 21 plus kit was used (Fast Track Diagnostics). This detects presence of 21 respiratory viruses which includes influenza A virus; influenza A(H1N1) virus (swine-lineage); influenza B virus; human rhinovirus; human coronaviruses NL63, 229E, OC43 and HKU1; human parainfluenza viruses 1, 2, 3 and 4; human metapneumoviruses A/B; human bocavirus; human respiratory syncytial viruses A/B; human adenovirus; enterovirus; human parechovirus; *Mycoplasma pneumoniae* and 4 bacterial species *S. aureus*; *C. pneumoniae*; *H. influenzae* B; *S. pneumonia*.

### Multiplex cytokine/chemokine assays

25 μl of serum was used for in magnetic bead-based multiplex chemokine and cytokine luminex assay as per the manufacturer’s instructions (Merck-Millipore). Assays were performed in duplicates along with quality controls. The amount of each analyte was estimated by standard curve generated using known amount of analyte provided in the assay kit. Assay plates were read using MAGPIX system and data was analysed by xPONENT software using five parametric logistic fit model.

### Cell Viability assay

Cell viability assay was performed using CellTiter-Glo® Assay kit (Promega) according to user manual. Briefly, 20,000 Calu-3 cells were seeded in 96 well plate and grown for 48 h. Cells were treated with zinc sulfate at indicated concentrations. After 24 h post treatment CellTiter-Glo reagent was added to the well and luminescence was measured by microplate reader (Synergy HT - BioTek).

### Labile zinc level measurement

Labile Zn levels in the cells were estimated as described before (17). Briefly, after incubation with zinc sulfate, cells were washed once with PBS, detached using trypsin and resuspended in staining medium (DMEM without phenol red supplemented and with 2 mM L-glutamine; Gibco) containing 2.5 μM ZinPyr-1 (ZP-1) (Santa Cruz) for 30 min at 37°C. Cells were washed using fluorescence-activated cell sorter (FACS) buffer (PBS containing 0.25% FBS), and samples were acquired in a FACS Canto II apparatus (Becton, Dickinson). The amounts of labile zinc present in the cells are presented as the mean fluorescence intensities of ZP-1.

### Immunofluoresence

VeroE6, Calu-3 (ATCC) and Caco-2 (ATCC) cells were seeded at 50,000 cells per well in 48-well plate. Calu-3 and Caco-2 cells were seeded 2 days prior whereas VeroE6 were seeded a day before infection. Cells were infected with SARS-CoV-2 THSTI-BL2010-D at 0.3, 1 and 3 MOI. After 1 h of adsorption, viral inoculum was removed. Cells were washed twice with PBS. Cells were kept in complete medium (10% FBS containing DMEM with penicillin, streptomycin and glutamine mix and non-essential amino acids). At 24 h pi, cells were washed twice with PBS and once with cold PBS and fixed by using ice cold methanol and kept at −20°C for 20 min. Methanol was removed, cells were washed once with cold PBS and then once with PBS at room temperature (RT), followed by blocking with 0.2% BSA-PBS for 10 min at RT. Cells were incubated with human chimeric SARS-CoV-2 S1 spike antibody (1: 100 dilution; Genescript – HC2001) in 0.2% BSA-PBS for 1h at RT. Cells were then washed three times with 0.2% BSA-PBS. This was followed by incubation with secondary antibody conjugated with anti-human Alexa flour 488 (1:500; Life Technologies–A11013) for 30 min at room temperature in dark. Cells were washed with PBS three times and stained with 4′,6-diamidino-2-phenylindole (DAPI) (Molecular probes) at 1:10,000 dilution for 10 min. Cells were washed with PBS and images were captured using Olympus DP80 microscope at 20X magnification. Images were processed by background subtraction using cellSens software. Bright field images were captured using Nikon Eclipse Ti-S at 10X magnification.

### Zinc treatment

Calu-3 cells (40-50K) were seeded in a 24-well plate and two days post-infection, cells were infected with 0.3 MOI of SARS-CoV-2. Cells were cultured in serum-free MEM containing indicated concentrations of zinc sulfate for 24 hours. Viral titers in the supernatant was measured by plaque assays as described above.

### Whole genome Sequencing and analysis

The protocol followed for the sample preparation, sequencing, and data analysis has been described earlier (18). In brief, double-stranded cDNA was synthesized from 50ng of total RNA from virus stocks for all the four plaque-purified SARS-CoV-2 isolates. The first strand of cDNA was synthesized using Superscript IV followed by RNA digestion with RNase H for second-strand synthesis using DNA Polymerase I Large fragment (Klenow fragment). 100ng of purified double-stranded cDNA was taken for forward using ARTIC tiling PCRprotocol (V3 primer pools) (https://artic.network/ncov-2019) (19). 200ng of each purified sample of multiplexed PCR amplicons obtained was taken for library preparation using Oxford Nanopore Technology (ONT). This included End Repair/dA tailing, Native Barcode Ligation, and Adapter Ligation of the PCR amplicons. 100ng of the pooled and purified library was sequenced using ONT’s MinION Mk1B platform (18).

Samples were base-called and demultiplexed using Guppy basecaller (version 3.5.2)(https://community.nanoporetech.com). Reads having phred quality score < 7 were discarded to filter the low-quality reads. The resulting fastq files were normalized by read length (300-500) and reads were aligned using Minimap2 (v2.17) (20) to the reference (GenBank: MN908947.3). Variants were called using Nanopolish(21) from the aligned reads and further creating consensus fasta using bcftools (v1.8). Finally, we used Nextclade (https://clades.nextstrain.org/) for the detection of clades from the consensus fastasequence. All the sequences are submitted to the GISAID database (https://www.gisaid.org/; GISAID EPI ISL - 454862).

### Phylogenetic analysis

The consensus genomes obtained from the whole genome sequencing of 10 Indian isolates and four plaque-purified Indian isolates along with 86 other global SARS-CoV-2 sequences were used for constructing the phylogeny. The sequences were selected from a diverse geographical location to represent the global distribution and spread of the virus. The sequences were obtained from the Global Initiative on Sharing All Influenza Data (GISAID) database. The fasta sequences were aligned using MUSCLE (22). A maximum-likehood Phylogeny was generated using FastTree 2.1.11 (23) with generalized time-reversible (GTR) substitution model and 1000 bootstrap replicates with default parameters. The tree was visualized and edited using ITOL (24).

### Trans-epithelial electrical resistance and confocal microscopy

Calu-3 cells were grown in air-liquid interface as described elsewhere (25). Briefly, Calu-3 cells were seeded at 30,000 on 3 μm pore size transwell inserts (Corning - 3415). Cells were maintained at liquid-liquid interface for 7 days and cells were further grown with 600 μl of complete medium only in the basolateral compartment. Media was replaced every alternate day. TEER was monitored at indicated time points using a chop stick electrode (Millipore). On day 21 post-seeding, cells were infected with SARS-CoV-2-THSTI-BL2010D as described. At 24 h pi, culture inserts were washed with cold 1X PBS and incubated with cold methanol at −20°C for 20 minutes. Cells were further incubated with IMF buffer (20 mM HEPES, pH 7.5, 0.1% Triton-X-100, 150 mM sodium chloride, 5 mM EDTA and 0.02% sodium azide as a preservative) for 5 min at room temperature (RT) and all further washes were performed with IMF buffer. Non-specific antibody binding sites were blocked by incubating cells with IMF buffer containing 2% normal goat serum for 10 min at RT. Membranes were cut out using a scalpel blade and transferred to a 48-well plate. Cells were washed three times followed by incubation with antibodies against ZO-1 (Becton Dickinson), Claudin-3 (Invitrogen), SARS-CoV-2 spike antibody in IMF buffer (20 mM HEPES pH 7.5, 0.1% Triton X-100, 150 mM NaCl, 5 mM EDTA, 0.02 % sodium azide) for 1 h at RT. Cells were washed followed by incubation with appropriate secondary antibodies tagged with Alexa fluor 488/568/633 (Molecular probes) for 30 min at RT by avoiding exposure to light. Cells were washed with IMF buffer three times and stained with DAPI at 1:10,000 dilution for 10 min. Cells were washed with PBS, mounted on glass slide using antifade solution (Molecular probes) and images were captured at 100X magnification using FV3000 confocal microscope (Olympus). Images were processed by background subtraction using cellSens software (Olympus).

### Data analysis

Data was analysed and final graphs were prepared using GraphPad Prism (Version 7.0e) software. All experiments were performed with two or more replicates and graphs have been prepared representing data from at least two independent experiments with n ≥ 6 unless otherwise indicated. Statistical significance was estimated by t-test (unpaired, non-parametric) using Mann-Whitney method.

## RESULTS

### Kinetics of viral load in COVID-19 patients with mild symptoms

11 patients who were positive for COVID-19 by RT-PCR were enrolled for the study after obtained informed consent. The median age of the patients was 32 (25% Percentile 24, 75% Percentile 45) and the mean duration of hospitalization was 14 days (Std. Dev. 4.034 days). Out of the 11 patients 6 were male and 5 were female. NP & OP swabs, saliva and serum samples collected for a maximum of 10 days. Most of the patients in this study manifested mild symptoms and were asymptomatic at the time of discharge. We measured daily viral load in the NP/OP samples by RT-PCR. For a subset of patients (n=5), we collected saliva samples early in the morning for estimation of viral load in saliva and compare with the NP/OP sample. Viral load in NP/OP swab samples peaked between day 2 to 4 in most patients and we observed a biphasic increase in viral RNA levels in few patients with a second peak on day 5 (Figure 1A-1F). The peak viral RNA levels ranged from 10^6^ to 10^10^ copies per ml of virus transport medium. Except for one patient, viral load in saliva correlated with viral load in VTM samples (Figure 1H-1K). In one patient, virus persisted in saliva till day 10 while the VTM sample was RT-PCR negative by day 6 of enrolment (Figure 1G). In most cases, the viral RNA was below the level of detection by RT-PCR by day 7-9 but two patients showed increasing trend in viral RNA levels although the last sample in these patients were on day 5 and day 7 (Figure 1D and 1H). These data suggest that the mild or asymptomatic patients harbour high viral load and salivary viral load can be a good alternate for the NP/OP swabs.

**Figure 1:**
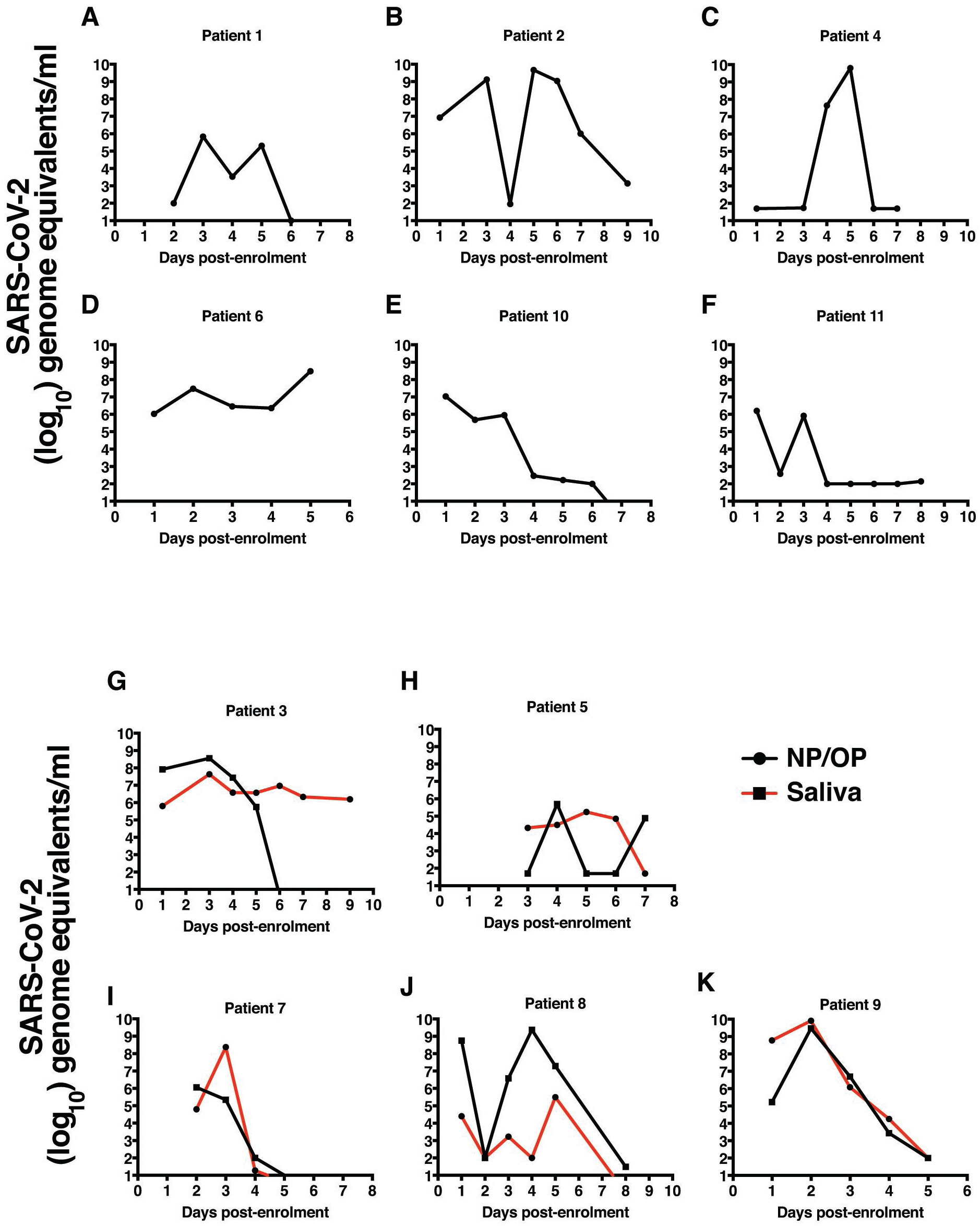
Kinetics of viral load in VTM and saliva samples of COVID-19 patients. A-F) SARS-CoV-2 RNA levels were measured by RT-PCR using total RNA isolated from VTM samples every day after enrolment. G-K) SARS-CoV-2 RNA levels were measured by RT-PCR using total RNA isolated from saliva and VTM samples every day after enrolment. Data points indicate geometric mean normalized to RNASe P levels which was used as endogenous controls for extraction efficiency.

### Kinetics of inflammatory mediators in mild/asymptomatic COVID-19 patients

As recent reports implicate both virus and host factors in severe COVID-19 disease, we were interested in estimating the levels of some of the inflammatory mediators along with interferons in the serum samples collected daily from mild COVID-19 patients to understand the correlates of recovery from disease. We measured 15 serum inflammatory mediators in each of the samples by multiplexing using luminex technology. The analytes detected are namely IFN-α2, IFN-γ, IL-1β, IL-4, IL6, IL-7, IL-8, IL10, IL12p40, IL12p70, IL17A, Monocyte Chemotactic Protein 1(MCP-1), MCP-3, TNF-α and gamma interferon inducible protein −10 (IP-10). We observed a consistent decrease in type I and type II during the acute phase of infection but both IFN-α2 and IFN-γ increased at later stages of infection coinciding with recovery and clearance of virus by day 8 of enrolment (Figure 2A). We next looked at the cytokines that influence T-cell functions namely the pro-inflammatory IL-12p40 homodimer or IL-12p70, the active IL-12 heterodimer secreted by dendritic cells and macrophages that promotes Th1 response and leads to IFN-γ and TNF-α production. We observed a clear increase in IL-12p70 levels from day 5 onwards suggesting priming of T cells by the innate immune system through IL-12p70 secretion (Figure 2B). This increase coincided with increased IFN-γ levels observed in the same samples (Figure 1A). IL-7, a cytokine produced by epithelial cells and DCs and promotes biogenesis and proliferation of lymphocytes also showed an increasing trend coinciding with IL-12p70.

**Figure 2:**
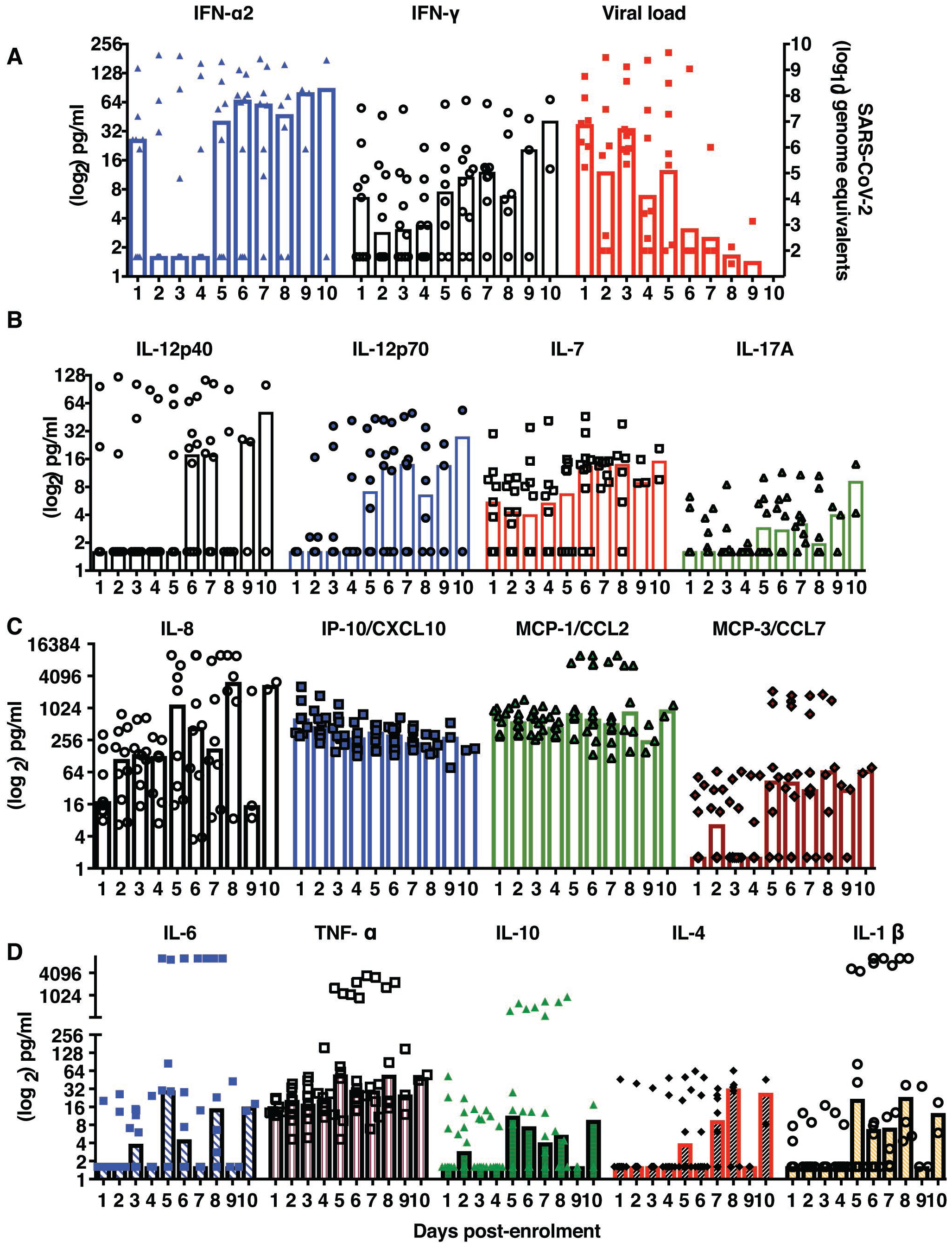
Kinetics of secreted inflammatory mediators in COVID-19 patients. A) Daily IFN-α, IFN-γ levels (on the left y-axis) were estimated from serum samples of COVID-19 patients using luminex assays and copy numbers of viral genome equivalents (on the right-y axis) was estimated from RNA isolated from VTM samples every day. B) The levels of indicated cytokines influencing T-cell functions were estimated from serum samples collected daily from COVID-19 patients. C) The levels of indicated chemokines suggestive of inflammation were estimated from serum samples collected daily from COVID-19 patients. D) The levels of indicated cytokines which are key determinants of inflammatory response were estimated from serum samples collected daily from COVID-19 patients. Columns indicate the median value with 95% CI.

However IL-17A levels did not show a significant change suggesting a more prominent role for Th1 responses in recovery in mild COVID-19 cases. Contrasting with the IFN responses, the levels of IL-8 was elevated at early stages of infection suggesting a key role for innate immune cells such as macrophages which produce IL-8 thus promoting neutrophil recruitment to the site of infection leading to resolution of infection. Interestingly, the levels of other two pro-inflammatory chemokines IP-10 and MCP-1 did not show any variation and MCP-3 levels were very heterogenous during the course of infection in any of the patients suggesting lack of severe inflammatory response in these patients (Figure 2C). IL-6, an acute phase reactant which is implicated in the severe manifestations of severe COVID-19 symptoms showed a moderate increase in multiple phases but TNF-α levels were unchanged in majority of the patients. The levels of IL-10, IL-4 and IL-1β showed an increasing trend in samples from day 5 - day 8 of infection suggesting the activation of T cells (Figure 2D). Overall our data suggests a prominent role for the innate immune response in clearance of virus infection/infected cells at early states of infection and priming of adaptive immune responses which probably lead to long-term protection from re-infection.

### Isolation and characterization of SARS-CoV-2

Increased viral loads have been associated with severe disease by previous reports (7, 15). We were interested in understanding whether the virus isolates from mild or asymptomatic infection have altered ability to evade immune response and cause cellular damage. In order to isolate and characterize an Indian isolate of SARS-CoV-2, we used NP/OP swab samples from six COVID-19 patients whose samples had a Ct value of 20 or less by the diagnostic RT-PCR. Vero E6 cells were inoculated with NP/OP swab samples and culture supernatant was harvested on day 2 post-infection when cytopathic effect was evident to generate passage 1 virus. The process was repeated two more times to generate passage 2 and passage 3 viruses (Supplementary Figure S1). One of the passage 3 isolate, THSTI-BL2010-D, was further plaque-purified by isolating four individual plaques (THSTI-BL2010-A through D) and propagating further one more time in Vero E6 cells to generate a large stock of passage 5 virus for all experiments (Figure 3A). All the plaque-purified virus showed CPE at 72 h pi. All the plaque-purified viruses were negative for 21 respiratory viruses and 4 bacterial species including Mycoplasma as assessed by RT-PCR indicating lack of any secondary pathogens. Next, we performed plaque assays to compare the plaque morphology of plaque-purified virus on VeroE6 cell-monolayer with SARS-CoV-2 USA-WA1/2020 isolate. Both the isolates developed plaques at 60 h pi, however, the plaque-purified isolate produced a more homogenous plaque size and distribution as compared to the USA-WA1/2020 isolate (Figure 3B).

**Figure 3:**
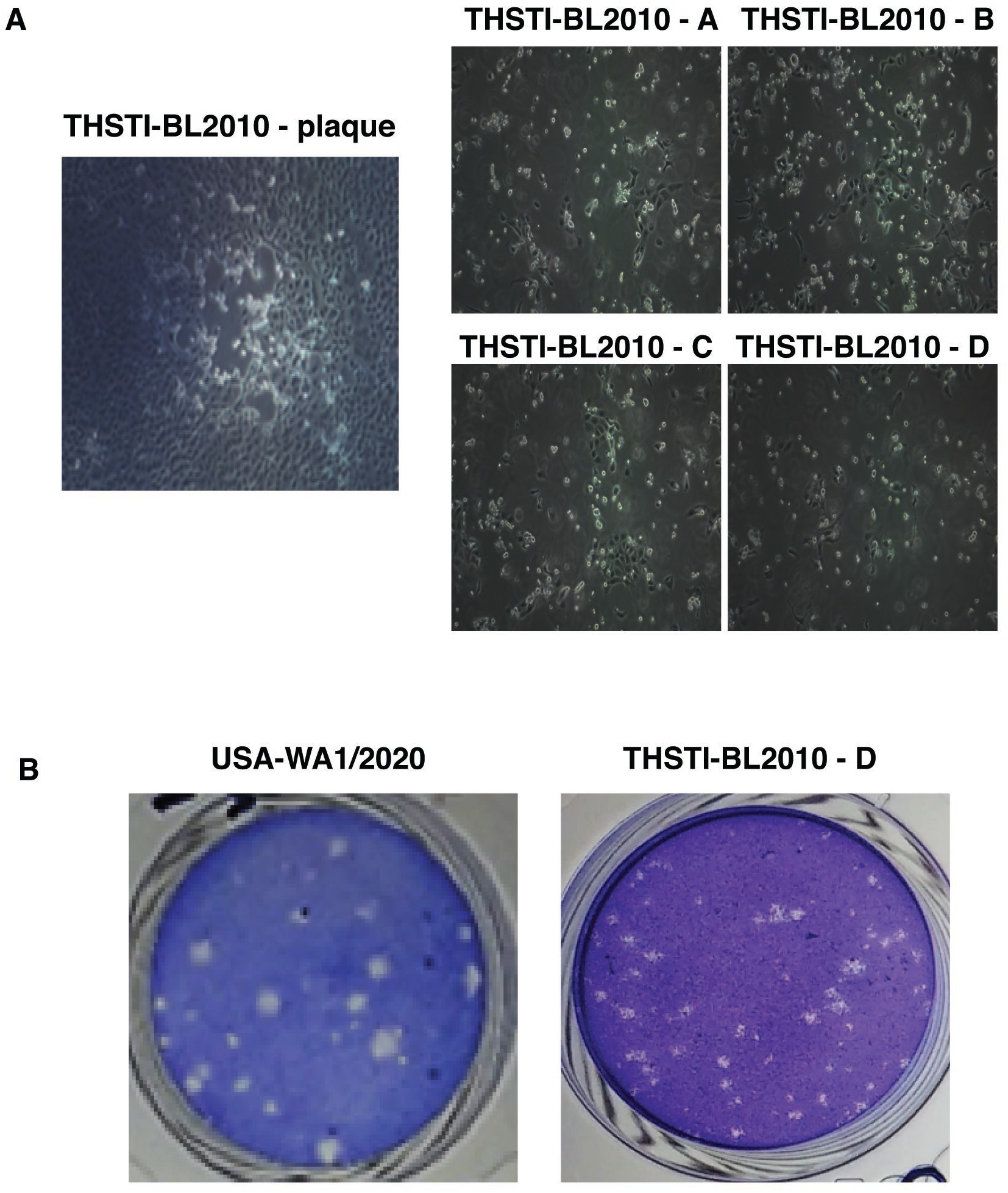
THSTI-BL-2010 plaque-purified isolates. A) Single plaque morphology of THSTI-BL-2010 at 72 h pi. Virus from four single plaques were propagated on VeroE6 monolayer separately. Cytopathic effect was evident at 72 h post-infection for all the four isolates. Images were captured at 20X magnification. B) VeroE6 cells were used for performing plaque assays with either the reference SARS-CoV-2 strain or THSTI-BL-2010D. Cells were fixed at 60 h pi and stained with crystal violet.

### SARS-CoV-2 infection in human epithelial cell lines

Infection of SARS-CoV-2 in human cell lines depends on the expression of its receptor angiotensin-converting enzyme 2 (ACE2) and the host protease TMPRSS2 (26). To confirm the infectivity of THSTI-BL2010-D in human cell lines, we infected Vero E6, Calu-3 and Caco-2 cells with increasing MOIs of the virus and stained for the presence of spike protein by immunofluorescence assay at 24 h pi. As expected, Vero E6 cells showed the maximum infection followed by Calu-3 cells and Caco-2 cells (Figure 4A). We next compared growth curve of SARS-CoV-2 Wuhan strain and THSTI-BL2010D in Vero E6 cells. Cells were infected with 0.1 MOI of both strains and supernatants were collected at 24, 48 and 72 h pi. Plaque assay results showed titres of Wuhan strain was a log higher than THSTI-BL2010D at 24 h pi whereas at 48 h pi virus titres with both strains were similar (Figure 4B). Virus titers showed a declining trend at 72 h pi most probably due to cytopathic effect.

**Figure 4:**
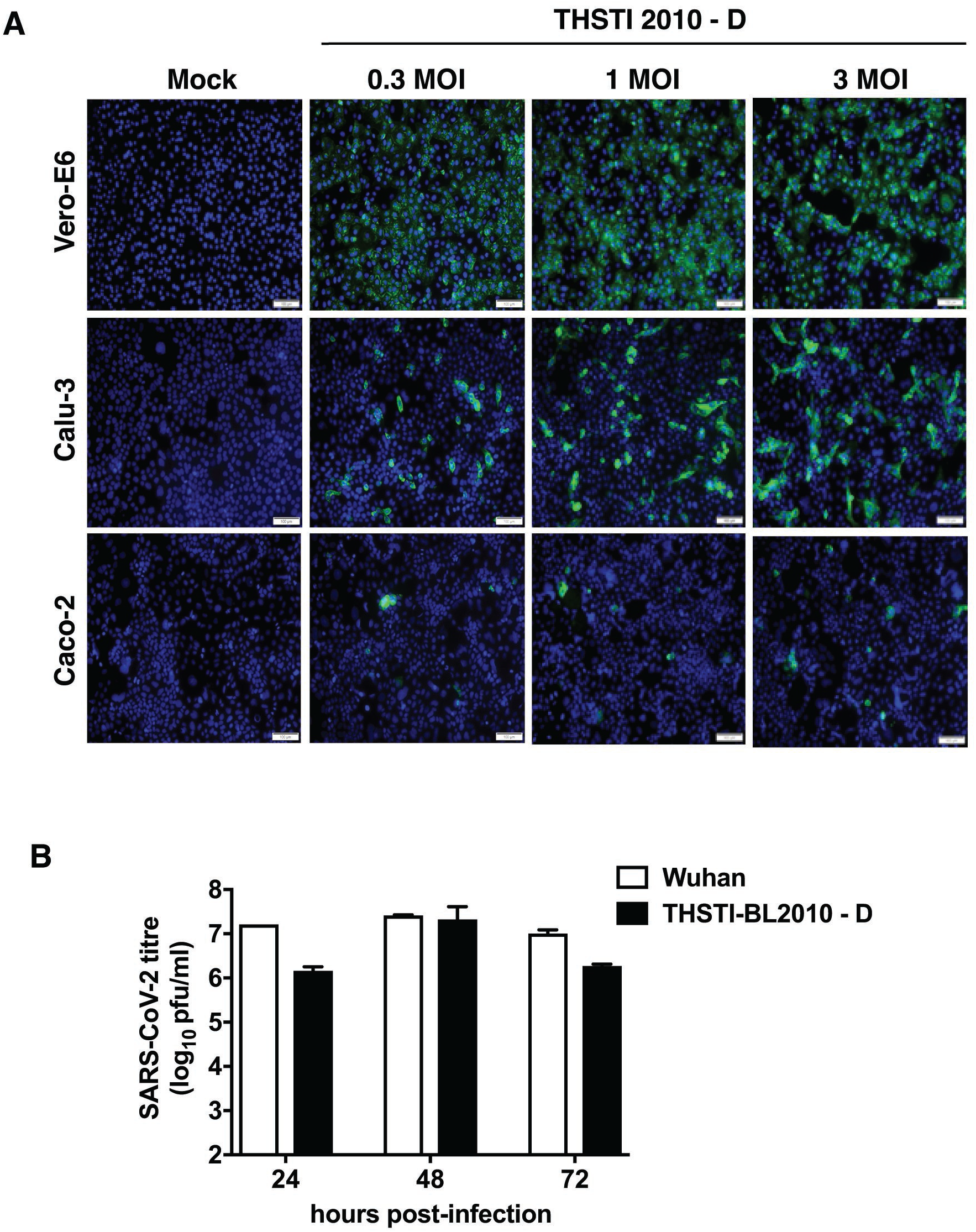
Infection of human cell lines. A) Vero E6, Calu-3 and Caco-2 cells were either mock-infected or infected with THSTI-2010-D at indicated MOI for 24 h. Cells were fixed in and stained with anti-spike protein antibody and probed with secondary antibody conjugated with Alexa Fluor 488. Images were captured using Olympus DP80 microscope. Scale bar is 100 μm. B) Growth curve of SARS-CoV-2 Wuhan and THSTI-BL2010D strain were compared on Vero E6 cells. Vero E6 cells were infected at 0.1 MOI either with Wuhan strain or THSTI BL2010D. Viral titers were measured at 24 h, 48 h and 72 h in culture supernatants (n=3).

### Whole genome sequencing

To further characterize the plaque-purified isolates at molecular level, we isolated RNA from the plaque-purified virus stocks and subjected to whole genome sequencing. All the four plaque-purified virus RNA samples showed similar changes in the nucleotide positions with most changes in the ORF1a region leading to five non-synonymous and one synonymous change in the protein sequence (Table 1). THSTI-BL-2010-C sample had one additional mutation G27806T resulting in a synonymous change in ORF7b. Two non-synonymous changes were also observed in ORF1b and N regions (Table 1). Interestingly, none of the Indian isolates in our study showed D614G mutations observed elsewhere which is associated with increased infectivity (27). The slippery heptanucleotide sequence and the pseudoknot structure were not disrupted in any of the samples as per the sequence alignment compared to the reference genome (Accession ID: MN908947.3). The phylogenetic analysis of the 100 representative SARS-CoV-2 sequences from around the world (Supplementary Table S1) revealed clustering of sequences into five major clades according to the GISAID nomenclature system (https://www.gisaid.org). Sequences from all the 14 genomes from our study were subjected to phylogenetic analysis and were assigned to clade 19A which along with the 19B clade forms the earliest clade which originated from Wuhan and was prevalent in Asia during the initial months of the outbreak (Figure 5). This suggests that the SARS-CoV-2 isolates reported in our study are genetically more similar to the reference Wuhan sequence (GenBank: MN908947.3).

**Table – 1.** List of mutations in plaque-purified isolates compared to the reference Wuhan strain (MN908947.3).

| **Virus** | **Nucleotide position** | **Base change** | **Open reading frame** | **Amino acid substitution** |
| --- | --- | --- | --- | --- |
| **THSTI-2010-**<br>**(A, B, C, D)** | 1227 | T→C | ORF1a | M321T |
|  | 1281 | C→T | ORF1a | A339V |
|  | 1758 | C→T | ORF1a | A498V |
|  | 4885 | T→C | ORF1a | Synonymous |
|  | 6312 | C→A | ORF1a | T2016K |
|  | 11083 | G→T | ORF1a | L3606F |
|  | 13730 | C→T | ORF1b | A88V |
|  | 23929 | C→T | Surface glycoprotein (S; Spike) | Synonymous |
|  | 28311 | C→T | Nucleocapsid phosphoprotein (N) | P13L |
| **THSTI-2010-C** | 27806 | G→T | ORF7b | Synonymous |

**Figure 5.**
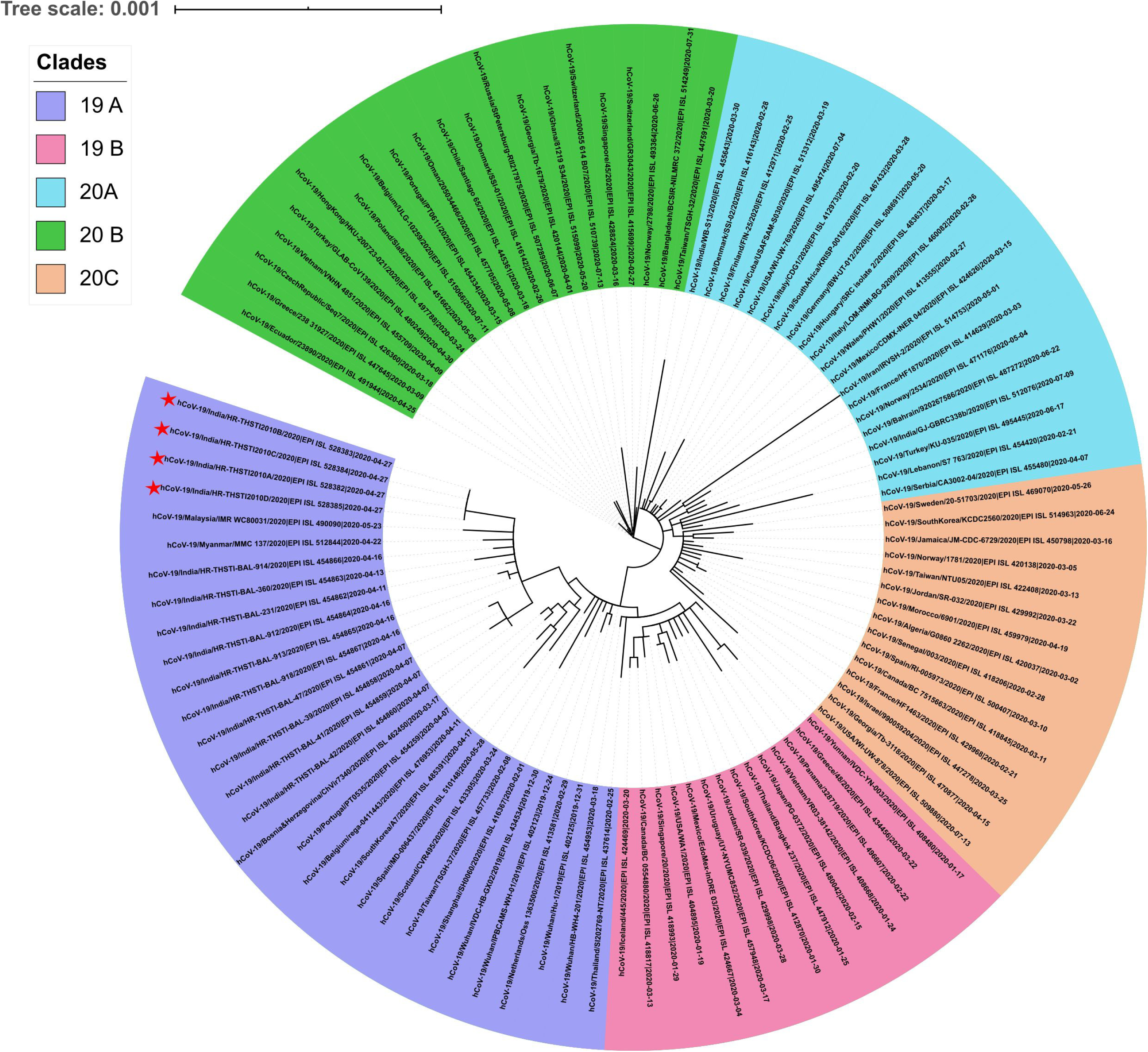
Maximum likelihood phylogeny for SARS CoV2 sequences (n=100). Indian isolates reported here falls under 19A clade. The sequence of plaque purified isolates are highlighted with symbol ‘star’. The GISAID clades are indicated as mentioned in the inset. The scale bar indicates the branch lengths measured in number of nucleotide substitution rate per site.

### SARS-CoV-2 infection disrupts epithelial barrier

We were further interested to investigate the effect of THSTI-BL-2010D isolate infection on epithelial barrier functions. Polarized Calu-3 cells grown on transwells were infected with SARS-CoV-2 at 0.3 MOI. TEER was measured at 24 h pi and virus titers in the apical and basolateral supernatants were estimated by plaque assays. We observed a 60% reduction in TEER values as compared to mock at 24 h pi indicating disruption of epithelial barriers as a result of cytopathic effect (Figure 6A). As expected, virus was present both in the apical and basolateral samples due to disruption of cellular junctions and polarity (Figure 6B). Cells were stained for virus spike protein, ZO-1 and claudin-3 antibodies to visualize infected cells and staining pattern of these representative junctional proteins. We observed disruption of both ZO-1 and claudin-3 staining in virus-infected cells but not in mock-infected cells (Figure 6C) suggesting that SARS-CoV-2 isolated from an mild/asymptomatic patient infects polarized lung epithelial cells and causes damage to epithelial barriers due to cytopathic effect.

**Figure 6:**
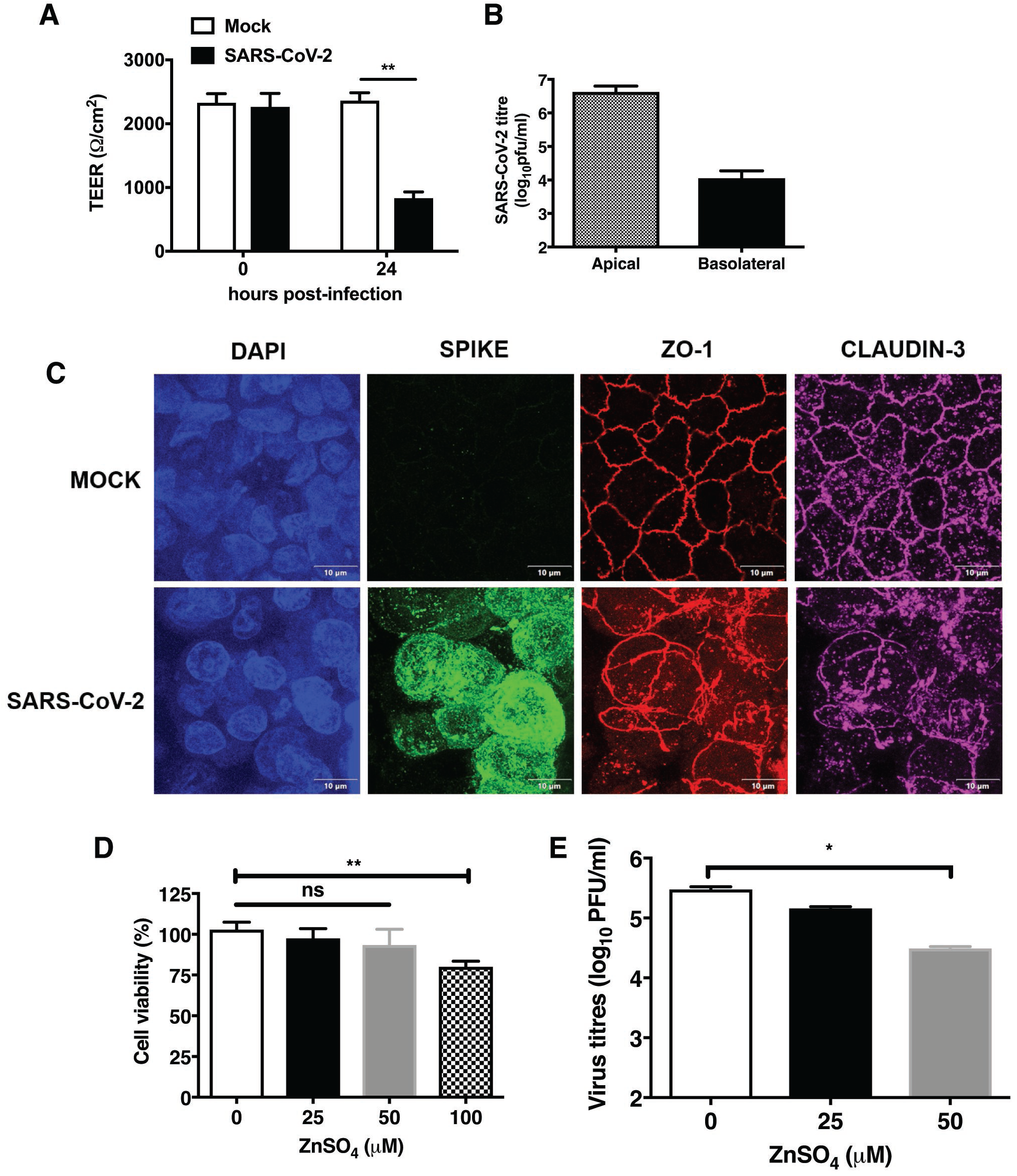
SARS-CoV-2 infection disrupts epithelial barrier integrity. A) Calu-3 cells grown on transwells at air-liquid interface as described in methods section. Polarized Calu-3 cells were infected with SARS-CoV-2 THSTI-BL-2010D at 0.3 MOI. TEER was measured at 24 h pi. TEER values were represented as ohm/cm^2^. B) Viral titres were determined at 24 h pi and represented as pfu/ml. C) Cells were fixed and stained with ZO-1 and Claudin-3 antibodies against cellular junctions and spike antibody against viral protein. Nuclei were stained with DAPI. Images were captured at 100X. Representative images of mock and SARS-CoV-2 infected cells are displayed. Scale bar is 10 μm. D) Calu-3 cells were treated with indicated concentrations of ZnSO_4_ for 24 h and cell viability was measured using CellTiter-Glo® Assay kit. E) Calu-3 cells were infected with SARS-CoV-2-THSTI-BL-2010D at 0.3 MOI. After viral adsorption, cells were treated with 25 and 50 μM of ZnSO_4_ or as mock treated control. Viral titres were determined as represented as pfu/ml. (n=3). Error bars represent mean ± SD. *p < 0.05, **p < 0.01.

### Zinc supplementation affects SARS-CoV-2 infection *in vitro*

Addition of zinc along with the zinc ionophore pyrithione, which stimulates zinc uptake, was shown to inhibit SARS-CoV infection in Vero E6 cells (28). We were interested to use our cell culture system to assess whether zinc has an inhibitory effect on SARS-CoV-2 infection. We first determined cytotoxicity and uptake of zinc in Calu-3 cells by treating cells with different concentration of ZnSO_4_ at 25, 50 and 100 μM for 24 h. We observed cytotoxicity beyond 50 μM ZnSO_4_ in Calu-3 cells (Figure 6D). Zinc uptake was determined by staining cells with zinc fluorophore, ZP-1, at 4 h post-treatment. Cells treated with 25 μM of ZnSO_4_ showed 25% increase in labile zinc levels whereas cells treated with 50 μM of ZnSO_4_ shows 50% increase in labile zinc levels as compared to mock-treated control suggesting that these cells are capable of zinc uptake (Supplementary Figure S2). Cells were infected with SARS-CoV-2 at 0.3 MOI and cultured in the presence of 25 and 50 μM of ZnSO_4_ for 24 h. At 24 h pi viral titres in the supernatant was determined by plaque assay. Zinc treatment showed over three-fold reduction in viral titers at 24 h pi as compared to untreated cells (Figure 6E) indicating the antiviral effect of zinc salts on SARS-CoV-2 infection in these cells.

## Discussion

Of all the coronavirus epidemic/pandemic of recent past, COVID-19 has had the maximum impact on global health and economy. Although most of the individuals are asymptomatic or with mild symptoms, the disease affects the elderly and people with co-morbidities the most indicating a critical role for innate immune response in limiting the viral load and spread. Most asymptomatic or individuals with mild symptoms harbour high viral loads in their respiratory tract and contribute to disease transmission. Previous report suggested reduced interferon response in severe patients as compared to mild/moderate cases 8-12 days after the onset of symptoms (7) and viral genetics was not found to correlate with severe disease (15). We focussed our efforts to monitor the kinetics of viral load and inflammatory mediators in the serum samples of hospitalized COVID-19 patients who displayed mild to no symptoms but were RT-PCR positive for SARS-CoV-2 during screening at the hospital or contact tracing. Most of our patients were enrolled within 48 hours of testing positive and are likely to be within the first week of exposure to the virus. In patients where we could detect viral RNA in saliva samples, the viral load in saliva correlated well with the same in VTM samples suggesting that saliva sample could replace NP/OP swabs which cause discomfort to patients during sample collection. Collection of saliva would also reduce the cost and time involved in sample collection for COVID-19 diagnosis and self-sampling by patients would reduce the risk of exposure for health care workers. The copy numbers of viral RNA, as estimated by quantifying the nucleocapsid gene region, varied by four orders magnitude suggesting multiple factors including the initial inoculum as a major determinants of viral load in the respiratory tract. Despite this huge variation in viral load, the clinical features and disease outcome did not vary as all patients were able to clear the virus and recover from the disease. This clearly suggests that a robust immune response in younger adults is capable to clearing the infection and age and imbalance in immune response could contribute to severe disease in older individuals and patients with co-morbidities. We observed that decrease in viral loads coincided with increase in IFN responses which is in agreement with observations made by other groups in *ex vivo*, cell culture and animal models where SARS-CoV-2 was shown to be a more potent inhibitor of IFN responses than SARS-CoV-1 (29, 30). Although our study does not have relative comparison between samples from different severities, we observed a consistent decrease in type I and type II during the acute phase of infection. Low serum interferons preceded respiratory failure in critical COVID-19 cases (7) whereas we observed an increase in both IFN-α and IFN-γ in all the patients who recovered and were discharged in our study clearly indicating a critical role for interferon responses in recovery from COVID-19. Increase in type I and II interferons indicate activation of T cells and NK cells in addition to the monocytes and macrophages which may contribute to the interferon levels. We have not assessed the levels of IFN-λ in these samples which is known to play a major role in antiviral response from epithelial cells (31). Our results show that there was moderate to no induction of pro-inflammatory cytokines or chemokines such as IL-6, TNF-α, IP-10 in any of these patients. We also observed, at later stages of infection, a trend showing increase in IL-10, IL-4 and IL-1β and IL-12p70 suggesting activation of T cells. Interestingly, IL-8, the primary chemoattractant for neutrophil recruitment has been shown to play an important role in acute respiratory distress syndrome (ARDS) and in recruitment of neutrophils to the site of injury in lungs in sepsis (32). We speculate that generation of high levels of IL-8 by the cells of innate immune system such as macrophages leading to activation of neutrophils which engage in viral clearance from the respiratory tract. Interestingly, neutrophils have been shown to play a crucial antiviral role in influenza A virus infection and in neurotropic coronavirus models such as mouse hepatitis virus (33). However, sustained levels of IL-8 may lead to ARDS observed in severe COVID-19 patients. Therefore, we hypothesize that regulation of IL-8 signalling and neutrophil activity could be an important determinant of clinical outcomes in COVID-19 patients.

An earlier study from India reported the isolation of SARS-CoV-2 from an Indian patient and its characterization (34). We got one step further and generate the first plaque-purified SARS-CoV-2 isolate from India. Plaque-purified SARS-CoV-2 revealed clear and homogeneous plaques on Vero E6 monolayers compared to the Wuhan strain where the plaques were very heterogeneous in size. As robust and consistent plaque reduction neutralization titers (PRNT) depend on high quality plaque assays, we believe our virus strain will help in establishing PRNT assays that could work reliably and consistently and help vaccine studies. Although SARS-CoV-2 primarily targets the respiratory system, *in vitro* and *in vivo* studies indicate broad tissue tropism (35). In contrast to a previous study which showed no difference between the permissibility of Calu-3 and Caco-2 cells for SARS-CoV-2 infection (35), the Indian SARS-CoV-2 isolate from our study infected Calu-3 cells better than Caco-2 cells. Interestingly, cell surface expressions of ACE2 on Caco-2 cells was shown to be higher than Calu-3 cells grown on trans-well inserts (36). Therefore, it would be interesting to investigate the difference in ACE2 expression under different growth conditions and passage history of cell lines to correlate with their susceptibility to infection by SARS-CoV-2. Finally, based on previous reports which demonstrate the inhibitory activity of zinc on coronavirus RNA polymerase (28) and the widely accepted beneficial role of zinc supplementation in respiratory infections (37), we show presence of excess zinc was inhibitory to SARS-CoV-2 in Calu-3 cells. As zinc is an acute-phase reactant which undergoes tissue redistribution during infection, our data suggests a beneficial effect of zinc supplementation in COVID-19 infection.

## Data Availability

All the data are presented within the manuscript and as supplemental information and is freely available.

## ACKNOWLEDGEMENTS

We thank Amresh Kumar Singh for technical support and all members of CCV lab and bioassay lab for their technical help and critical inputs. We thank all the patients who consented to participate in the study.

## AUTHOR CONTRIBUTIONS

AA, SG, SK, JW, NAK, AP, AK, VA, JSV and RP performed experiments and analyzed the data. NV, AD and AKP coordinated the study at clinical site and contributed reagents. GRM conceived the study, designed the experiments, and analyzed data. AA, SG, SK, JW and GRM wrote the manuscript. All authors have reviewed and approved the final version of the manuscript.

## SUPPLEMENTARY INFORMATION

This article has supplementary information.

## SUPPLEMENTARY FIGURE LEGENDS

**Figure S1: SARS-CoV-2 Indian isolates**. Vero E6 cells were infected with nasopharyngeal swab samples for 1 h and cultured in growth medium thereafter. CPE was evident at 48 h post-infection. Images were captured using Nikon Eclipse Ti-S microscope at 20X magnification.

**Figure S2: Zinc uptake in Calu-3 cells**. Calu-3 cells after incubation with indicated concentrations of zinc sulfate for 4 h and labile zinc levels were measured by staining with ZinPyr-1 and flow cytometry. The relative change in the mean fluorescence intensities of ZP-1 is shown in the graph. Error bars represent Mean with SD.

