## Supplementary figures and images for "Kinetics of viral load, immunological mediators and characterization of a SARS-CoV-2 isolate in mild COVID-19 patients during acute phase of infection"

### Supplementary Figures S1 and S2

**Figure S1**

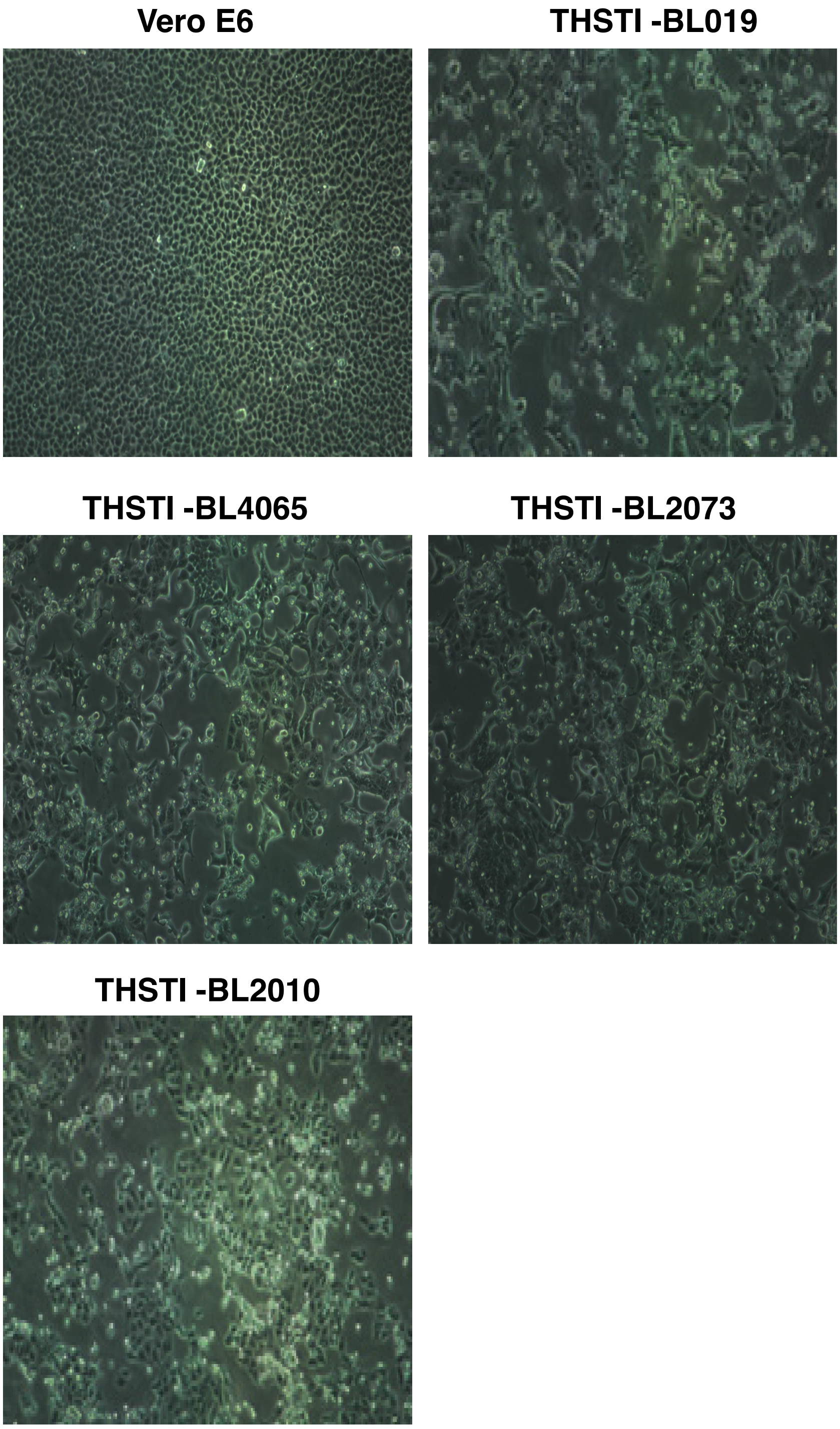

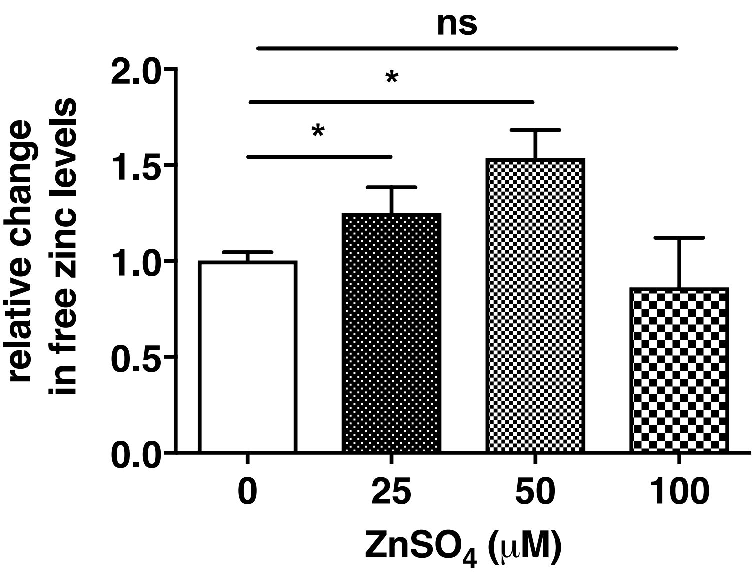
