## Supplementary Table S1 for "Kinetics of viral load, immunological mediators and characterization of a SARS-CoV-2 isolate in mild COVID-19 patients during acute phase of infection"

**Supplementary Table S1**: List of sequences used in constructing phylogenetic tree.

| **SARS-CoV-2 isolate** | **GISAID ID** | **Collection date** |
| --- | --- | --- |
| Algeria/G0860_2262/2020 | EPI_ISL_420037 | 2020/03/02 |
| Bahrain/920267586/2020 | EPI_ISL_487272 | 2020/06/22 |
| Bangladesh/BCSIR-NILMRC_372/2020 | EPI_ISL_514249 | 2020/07/31 |
| Belgium/rega-0411443/2020 | EPI_ISL_476953 | 2020/04/11 |
| Belgium/ULG-10259/2020 | EPI_ISL_515066 | 2020/07/11 |
| Bosnia & Herzegovina/ChVir7340/2020 | EPI_ISL_462450 | 2020/03/17 |
| Canada/BC_0554880/2020 | EPI_ISL_418817 | 2020/03/13 |
| Canada/BC_7515663/2020 | EPI_ISL_418845 | 2020/03/11 |
| Chile/Santiago_65/2020 | EPI_ISL_445361 | 2020/03/18 |
| Cuba/USAFSAM-S030/2020 | EPI_ISL_513312 | 2020/03/19 |
| Czech Republic/Seq7/2020 | EPI_ISL_426360 | 2020/03/18 |
| Denmark/SSI-01/2020 | EPI_ISL_416142 | 2020/02/26 |
| Denmark/SSI-02/2020 | EPI_ISL_416143 | 2020/02/28 |
| Ecuador/23890/2020 | EPI_ISL_491944 | 2020/04/25 |
| Finland/FIN-25/2020 | EPI_ISL_412971 | 2020/02/25 |
| France/HF1463/2020 | EPI_ISL_429968 | 2020/02/21 |
| France/HF1870/2020 | EPI_ISL_414629 | 2020/03/03 |
| Georgia/Tb-1679/2020 | EPI_ISL_420144 | 2020/04/01 |
| Georgia/Tb-3118/2020 | EPI_ISL_470877 | 2020/04/15 |
| Germany/BW-UT-012/2020 | EPI_ISL_508691 | 2020/05/20 |
| Ghana/81219_S34/2020 | EPI_ISL_515099 | 2020/05/20 |
| Greece/238_31927/2020 | EPI_ISL_447645 | 2020/03/09 |
| Greece/48/2020 | EPI_ISL_434456 | 2020/03/22 |
| Hong Kong/HKU-200723-021/2020 | EPI_ISL_497788 | 2020/03/24 |
| Hungary/SRC_isolate_2/2020 | EPI_ISL_483637 | 2020/03/17 |
| Iceland/445/2020 | EPI_ISL_424469 | 2020/03/20 |
| India/GJ-GBRC338b/2020 | EPI_ISL_512076 | 2020/07/09 |
| India/HR-THSTI2010A/2020 | EPI_ISL_528382 | 2020/04/27 |
| India/HR-THSTI2010B/2020 | EPI_ISL_528383 | 2020/04/27 |
| India/HR-THSTI2010C/2020 | EPI_ISL_528384 | 2020/04/27 |
| India/HR-THSTI2010D/2020 | EPI_ISL_528385 | 2020/04/27 |
| India/HR-THSTI-BAL-231/2020 | EPI_ISL_454862 | 2020/04/11 |
| India/HR-THSTI-BAL-360/2020 | EPI_ISL_454863 | 2020/04/13 |
| India/HR-THSTI-BAL-39/2020 | EPI_ISL_454858 | 2020/04/07 |
| India/HR-THSTI-BAL-41/2020 | EPI_ISL_454859 | 2020/04/07 |
| India/HR-THSTI-BAL-42/2020 | EPI_ISL_454860 | 2020/04/07 |
| India/HR-THSTI-BAL-47/2020 | EPI_ISL_454861 | 2020/04/07 |
| India/HR-THSTI-BAL-912/2020 | EPI_ISL_454864 | 2020/04/16 |
| India/HR-THSTI-BAL-913/2020 | EPI_ISL_454865 | 2020/04/16 |
| India/HR-THSTI-BAL-914/2020 | EPI_ISL_454866 | 2020/04/16 |
| India/HR-THSTI-BAL-918/2020 | EPI_ISL_454867 | 2020/04/16 |
| India/WB-S13/2020 | EPI_ISL_455643 | 2020/03/30 |
| Iran/IRVSH-2/2020 | EPI_ISL_514753 | 2020/05/01 |
| Israel/990059204/2020 | EPI_ISL_447278 | 2020/03/25 |
| Italy/CDG1/2020 | EPI_ISL_412973 | 2020/02/20 |
| Italy/LOM-INMI-BG-9209/2020 | EPI_ISL_460082 | 2020/02/26 |
| Jamaica/JM-CDC-6729/2020 | EPI_ISL_450798 | 2020/03/16 |
| Japan/PG-0372/2020 | EPI_ISL_480042 | 2020/02/15 |
| Jordan/SR-032/2020 | EPI_ISL_429992 | 2020/03/22 |
| Jordan/SR-039/2020 | EPI_ISL_429998 | 2020/03/28 |
| Lebanon/S7_763/2020 | EPI_ISL_454420 | 2020/02/21 |
| Malaysia/IMR_WC80031/2020 | EPI_ISL_490090 | 2020/05/23 |
| Mexico/CDMX-INER_04/2020 | EPI_ISL_424626 | 2020/03/15 |
| Mexico/EdoMex-InDRE_03/2020 | EPI_ISL_424667 | 2020/03/04 |
| Morocco/6901/2020 | EPI_ISL_459979 | 2020/04/19 |
| Myanmar/MMC_137/2020 | EPI_ISL_512844 | 2020/04/22 |
| Netherlands/Oss_1363500/2020 | EPI_ISL_413581 | 2020/02/29 |
| Norway/1781/2020 | EPI_ISL_420138 | 2020/03/05 |
| Norway/2534/2020 | EPI_ISL_471176 | 2020/05/04 |
| Norway/2798/2020 | EPI_ISL_493364 | 2020/06/26 |
| Oman/205034466/2020 | EPI_ISL_457705 | 2020/05/08 |
| Panama/328719/2020 | EPI_ISL_496607 | 2020/02/22 |
| Poland/Sla8/2020 | EPI_ISL_451662 | 2020/05/05 |
| Portugal/PT0535/2020 | EPI_ISL_454259 | 2020/04/07 |
| Portugal/PT0611/2020 | EPI_ISL_454334 | 2020/03/15 |
| Russia/StPetersburg-RII21797S/2020 | EPI_ISL_507289 | 2020/06/07 |
| Scotland/CVR495/2020 | EPI_ISL_433305 | 2020/03/24 |
| Senegal/003/2020 | EPI_ISL_418206 | 2020/02/28 |
| Serbia/CA3002-04/2020 | EPI_ISL_455480 | 2020/04/07 |
| Shanghai/SH0060/2020 | EPI_ISL_416367 | 2020/02/01 |
| Singapore/20/2020 | EPI_ISL_418993 | 2020/01/29 |
| Singapore/45/2020 | EPI_ISL_428824 | 2020/03/16 |
| South Africa/KRISP-0016/2020 | EPI_ISL_467432 | 2020/03/28 |
| South Korea/A7/2020 | EPI_ISL_485391 | 2020/04/17 |
| South Korea/KCDC06/2020 | EPI_ISL_412870 | 2020/01/30 |
| South Korea/KCDC2560/2020 | EPI_ISL_514963 | 2020/06/24 |
| Spain/MD-006437/2020 | EPI_ISL_510148 | 2020/05/28 |
| Spain/RI-005973/2020 | EPI_ISL_500407 | 2020/03/10 |
| Sweden/20-51703/2020 | EPI_ISL_469070 | 2020/05/26 |
| Switzerland/200055_614_B07/2020 | EPI_ISL_510739 | 2020/07/13 |
| Switzerland/GR3043/2020 | EPI_ISL_415699 | 2020/02/27 |
| Taiwan/NTU05/2020 | EPI_ISL_422408 | 2020/03/13 |
| Taiwan/TSGH-32/2020 | EPI_ISL_447591 | 2020/03/20 |
| Taiwan/TSGH-37/2020 | EPI_ISL_457733 | 2020/02/08 |
| Thailand/Bangkok_237/2020 | EPI_ISL_447912 | 2020/01/25 |
| Thailand/SI202769-NT/2020 | EPI_ISL_437614 | 2020/02/25 |
| Turkey/GLAB-CoV139/2020 | EPI_ISL_480249 | 2020/04/30 |
| Turkey/KU-035/2020 | EPI_ISL_495445 | 2020/06/17 |
| Uruguay/UY-NYUMC852/2020 | EPI_ISL_457948 | 2020/03/17 |
| USA/WA1/2020 | EPI_ISL_404895 | 2020/01/19 |
| USA/WI-UW-769/2020 | EPI_ISL_495474 | 2020/07/04 |
| USA/WI-UW-878/2020 | EPI_ISL_509880 | 2020/07/13 |
| Vietnam/VNHN_4851/2020 | EPI_ISL_455709 | 2020/04/09 |
| Vietnam/VR03-38142/2020 | EPI_ISL_408668 | 2020/01/24 |
| Wales/PHW1/2020 | EPI_ISL_413555 | 2020/02/27 |
| Wuhan/HB-WH4-201/2020 | EPI_ISL_454953 | 2020/03/18 |
| Wuhan/Hu-1/2019 | EPI_ISL_402125 | 2019/12/31 |
| Wuhan/IPBCAMS-WH-01/2019 | EPI_ISL_402123 | 2019/12/24 |
| Wuhan/IVDC-HB-GX02/2019 | EPI_ISL_434534 | 2019/12/30 |
| Yunnan/IVDC-YN-003/2020 | EPI_ISL_408480 | 2020/01/17 |
